# Urate, blood pressure and cardiovascular disease: updated evidence from Mendelian randomization and meta-analysis of clinical trials

**DOI:** 10.1101/2019.12.11.19014472

**Authors:** Dipender Gill, Alan C. Cameron, Stephen Burgess, Xue Li, Daniel J. Doherty, Ville Karhunen, Azmil H Abdul-Rahim, Martin Taylor-Rowan, Verena Zuber, Philip S. Tsao, Derek Klarin, VA Million Veteran Program, Evangelos Evangelou, Paul Elliott, Scott M. Damrauer, Terence J Quinn, Abbas Dehghan, Evropi Theodoratou, Jesse Dawson, Ioanna Tzoulaki

**Affiliations:** Department of Epidemiology and Biostatistics, School of Public Health, Imperial College London, London, UK; Institute of Cardiovascular & Medical Sciences, University of Glasgow, Glasgow, UK; MRC Biostatistics Unit, Cambridge Institute of Public Health, Cambridge, UK; Cardiovascular Epidemiology Unit, Department of Public Health and Primary Care, University of Cambridge, Cambridge, UK; Centre for Global Health, Usher Institute, University of Edinburgh, Edinburgh, UK; Institute of Neuroscience and Psychology, University of Glasgow, Glasgow, UK; VA Palo Alto Health Care System, California, USA; Department of Medicine, Stanford University School of Medicine, California, USA; Malcom Randall VA Medical Center, Gainesville, Florida, USA; Center for Genomic Medicine, Massachusetts General Hospital, Harvard Medical School, Massachusetts, USA; Program in Medical and Population Genetics, Broad Institute of MIT and Harvard, Massachusetts, USA; Division of Vascular Surgery and Endovascular Therapy, University of Florida School of Medicine, Gainesville, Florida, USA; Department of Hygiene and Epidemiology, University of Ioannina Medical School, Ioannina, Greece; MRC Centre for Environment and Health, School of Public Health, Imperial College London, London, UK; UK Dementia Research Institute at Imperial College London, London, UK; Imperial Biomedical Research Centre, Imperial College London and Imperial College NHS Healthcare Trust, London, UK; Health Data Research UK-London, London, UK; Corporal Michael J. Crescenz VA Medical Center, Philadelphia, USA; Department of Surgery, University of Pennsylvania Perelman School of Medicine, Philadelphia, USA; Edinburgh Cancer Research Centre, Institute of Genetics and Molecular Medicine, University of Edinburgh, Edinburgh, United Kingdom

**Author notes:** These authors contributed equally and are joint first (Dipender Gill, Alan C. Cameron). These authors contributed equally and are joint last (Evropi Theodoratou, Jesse Dawson, Ioanna Tzoulaki). **Corresponding author** Dr Dipender Gill, Department of Epidemiology and Biostatistics, School of Public Health, Medical School Building, St, Mary’s Hospital, Imperial College London, United Kingdom, W2 1PG.

**Keywords:** urate, cardiovascular disease, blood pressure, Mendelian randomization, systematic review

## Abstract

**Aims:** To investigate the effect of serum urate on blood pressure and cardiovascular disease (CVD) risk by updating evidence from Mendelian randomization (MR) analysis and meta-analysis of randomized controlled trials (RCTs).

**Methods and Results:** Using recently available data from the Million Veterans Program and UK Biobank, the main MR analyses showed that every 1-standard deviation increase in genetically predicted serum urate was associated with an increased risk of coronary heart disease (odds ratio 1.19, 95% confidence interval 1.10-1.30, *P*=4×10^−5^), peripheral artery disease (1.12, 95%CI 1.03 to 1.21, *P*=9×10^−3^), and stroke (1.11, 95%CI 1.05 to 1.18, *P*=2×10^−4^). In MR mediation analyses, SBP was estimated to mediate approximately one third the effect of urate on CVD risk. Systematic review and meta-analysis of RCTs showed a favorable effect of urate-lowering treatment on SBP (mean difference -2.55mmHg, 95%CI -4.06 to -1.05, *P*=1×10^−3^) and major adverse cardiovascular events (MACE) in those with previous CVD (OR 0.40, 95%CI 0.22 to 0.73, *P*=3×10^−3^), but no significant effect on MACE in all individuals (OR 0.73, 95%CI 0.48 to 1.09, *P*=0.12).

**Conclusion:** MR and clinical trial data support an effect of higher serum urate on increasing blood pressure, which may mediate a consequent effect on CVD risk. High-quality trials are necessary to provide definitive evidence on the specific clinical contexts where urate-lowering may be of cardiovascular benefit.

## Introduction

Urate is a breakdown product of purine metabolism. Its raised levels have been associated with a number of adverse health outcomes including gout, hypertension and cardiovascular disease (CVD) (1). However, it remains unclear whether these associations represent causal effects (2, 3). The relationship between serum urate, obesity, diet and other cardiovascular risk factors raises considerable potential for confounding and reverse causation (4).

Pre-clinical studies support a causal role for urate in hypertension (5). Randomized controlled trial (RCT) data have shown that both allopurinol and probenecid reduce systolic blood pressure (SBP) in hyperuricemic adolescents (1, 6, 7). Pooling of all available trial data can offer more precise effect estimates for the effect of urate-lowering therapy on blood pressure (8). As elevated blood pressure is a risk factor for CVD, it is important to clarify any role of SBP in mediating an effect of urate on cardiovascular outcomes.

The genetic determinants of serum urate levels have been increasingly well-characterized (4). This has made it possible to identify better instruments for Mendelian randomization (MR) analyses investigating the effect of genetically predicted serum urate on cardiovascular outcomes than in previous efforts (3, 9-11). The use of variants randomly allocated at conception to proxy the effect of modifying serum urate means that MR is less susceptible to the environmental confounding, measurement error and reverse causation bias that can limit causal inference in traditional epidemiological approaches.

The aim of the current study was to perform MR analyses investigating the effect of genetically predicted urate levels on SBP and CVD risk using contemporary data, and compare findings with results obtained from updated systematic review and meta-analysis of urate-lowering RCTs.

## Methods

### Two-sample Mendelian randomization

The MR analyses have been reported as per the STROBE-MR guidelines (Supplementary Checklist 1) (12).

#### Genetic association estimates

Genetic association estimates for serum urate in two-sample MR were obtained by using PLINK software to meta-analyze summary data from GWAS analyses of 110,347 European-ancestry individuals and 343,836 White British UK Biobank participants respectively (13-15). Urate estimates in MR analyses are provided per 1-standard deviation (SD), which corresponds to 80.3µmol/L. For consideration of SBP as a mediator in the two-sample multivariable MR, genetic association estimates were obtained from a GWAS of 317,195 White British UK Biobank participants, where SBP was measured using automated readings with correction made for any anti-hypertensive drug use by adding 10mmHg to the measured reading (16). In contrast, when investigating SBP as an outcome, genetic association estimates were obtained from the International Consortium for Blood Pressure (ICBP) GWAS analysis of 287,245 European-ancestry individuals (excluding UK Biobank participants) (17). A different population was considered when studying SBP as an outcome to avoid overlap with the UK Biobank participants used to obtain genetic association estimates for urate, as this can bias MR estimates (18). SBP estimates are provided per 1-SD increase, which corresponds to 18.6mmHg. Genetic association estimates for CHD were obtained from the CARDIoGRAMplusC4D Consortium 1000G multi-ethnic GWAS (77% European-ancestry) of 60,801 cases and 123,504 controls, with a broad and inclusive definition of CHD applied (19). Genetic association estimates for PAD were obtained from the Million Veterans Program (MVP) multi-ethnic (72% European-ancestry) GWAS of 31,307 cases and 211,753 controls, with case definitions made using hospital diagnosis and procedure codes (20). Genetic association estimates for stroke were obtained from the MEGASTROKE multi-ethnic (86% European-ancestry) GWAS of 67,162 cases and 454,450 controls (21), with the stroke definition including both ischemic and hemorrhagic etiologies.

#### Genetic variants used as instruments

Genetic instruments for the two-sample MR were identified as single-nucleotide polymorphisms (SNPs) that were associated with urate (or SBP, in mediation analyses) at genome-wide significance (P<5×10^−8^) and were in pair-wise linkage disequilibrium (LD) with *r*^2^<0.001. Clumping was performed using the TwoSampleMR package of R (22). For univariable MR, instrument strength was estimated using the *F*-statistic, with variance in the exposure explained assessed using the *R*^2^ value (23).

#### Statistical analysis

In all analyses, SNPs were aligned by their effect alleles and no additional consideration was made for palindromic variants. Two-sample MR analyses were performed to investigate the effect of genetically predicted serum urate on CHD, PAD, stroke and SBP respectively. A Bonferroni threshold (*P*<0.01) that corrected for multiple testing related to the four outcomes was used to ascertain statistical significance in the main analysis. Inverse-variance weighted MR was used in the main analysis, with the simple median (24), contamination-mixture method (25), Egger (26), PRESSO (27) and multivariable MR (28) (only for CVD outcomes) sensitivity analyses used to explore the robustness of the findings to potential pleiotropic of the genetic variants. Given the previously demonstrated overlap in the genetic determinants of urate with other metabolic traits (4), the multivariable MR sensitivity analysis adjusted for genetic associations of the instruments with body mass index (BMI), estimated glomerular filtration rate, type 2 diabetes mellitus, serum low-density lipoprotein cholesterol, serum high-density lipoprotein cholesterol and serum triglycerides together in the same model. Such multivariable MR was not performed when considering SBP as an outcome, due to population overlap with the cohorts used to obtain genetic association estimates for the metabolic exposures (18). In MR mediation analyses, multivariable MR was applied in the two-sample setting to adjust for the genetic association of the instruments with SBP, and network MR was used to estimate the proportion of the total effect of urate on each cardiovascular outcome that is mediated through SBP (29). Standard errors were estimated using the propagation of error method. Further details on these analyses are provided in the Supplementary Methods.

Genetic association estimates, instruments and statistical analyses for the one-sample MR are detailed in the Supplementary Methods.

### Systematic review and meta-analysis of randomized controlled trials

The systematic review was conducted in accordance with the recommendations from the Preferred Reporting Items for Systematic Reviews and Meta-analyze (PRISMA) statement (Supplementary Checklist 2) (30). The study protocol was registered and published in the International Prospective Register of Systematic Reviews (PROSPERO: CRD42020164589).

#### Inclusion and exclusion criteria

RCTs of pharmacological urate-lowering versus placebo or no treatment of duration ≥ 28 days in patients ≥ 18 years of age were eligible. Studies with co-intervention that was inconsistent between intervention and control groups, or with mixed experimental groups were excluded.

#### Data sources and searches

We conducted a systematic search of multidisciplinary databases across four electronic platforms: Medline (OVID), EMBASE (OVID), Web of Science (Thomson Reuters) and Cochrane Library (Cochrane). Studies published from 1 January 2016 to 30 September 2019 were considered. The literature search terms matched those used in a previous systematic review (Supplementary Methods) (7). No language restrictions were applied. Reference lists of retrieved articles were reviewed to identify additional relevant articles. Data were included from studies in previous systematic reviews investigating the effects of urate lowering therapy on SBP (searches until 29 June 2016) (8), and CVD (searches until 30 December 2016) (7).

#### Data extraction

Study details and results were extracted using a pre-designed template. In the case of cross-over trials, only data from the first study period (before cross-over) were used. Risk of bias was assessed in accordance with the Cochrane guidelines (31). All aspects of database search, study selection, data extraction and risk of bias assessment were performed independently by two investigators (ACC and DJD), with arbitration to a third investigator (AHA-R) as necessary.

#### Outcomes

The outcomes considered in systematic review and meta-analysis of RCTs were SBP and risk of major adverse cardiovascular event (MACE; cardiovascular death, non-fatal myocardial infarction, unstable angina requiring urgent revascularization, or non-fatal stroke). In secondary analysis, we also considered MACE in patients with prior CVD, as this would likely represent a higher risk population with greater statistical power.

#### Statistical analysis

Analysis of extracted data was based on modified intention-to-treat (considering patients who received at least one dose of the allocated treatment) or intention-to-treat results. If not reported, the mean change in SBP pre-and post-treatment was calculated and the SD was imputed using the propagation of error method using a correlation coefficient that was derived from the largest study in the meta-analysis that reported SD for SBP values at baseline, at end of the study and for the change in SBP (31, 32). The measure of effect size was the difference in mean SBP, defined as the mean difference between patients treated with urate lowering therapy versus control. A negative value of the difference in mean SBP indicates a greater reduction of SBP in the urate lowering therapy group as compared to the control group. Overall CVD risk estimates are described in terms of odds-ratio (OR).

Random-effects inverse-variance weighted meta-analysis was used to pool estimates from different studies, and study heterogeneity was assessed using the I-squared (I^2^) statistic. Sensitivity analyses were conducted including only studies at low risk of bias. For the analysis considering SBP as an outcome, meta-regression was performed to assess the effect of baseline SBP on treatment response. Analyses were conducted using Comprehensive Meta-Analysis software version 3 (Biostat, USA).

### Ethical approval, data availability and reporting

All data used in this work are either publicly accessible or available on request from their original studies, which obtained appropriate patient consent and ethical approval. The UK Biobank data were accessed through application 236. All results generated in this work are presented in either the manuscript or its supplementary files.

## Results

### Mendelian randomization

The instruments for urate that were used in the two-sample MR are presented in Supplementary Table 2. The main IVW MR showed that higher genetically predicted serum urate levels were associated with an increased risk of CHD, with odds ratio (OR) per 1-SD increase in genetically predicted urate 1.19, 95% confidence interval (95%CI) 1.10-1.30, *P*=4×10^−5^. Consistent results were obtained in all MR sensitivity analyses except MR-Egger, which had wide 95%CIs (Figure 1). Higher genetically predicted serum urate was also associated with increased risk of both PAD (OR 1.12, 95%CI 1.03-1.21, *P*=9×10^−3^) and stroke (OR 1.11, 95%CI 1.05-1.18, *P*=2×10^−4^) in the main IVW analysis, with similar results obtained in all MR sensitivity analyses except MR-Egger, which again had wide 95%CIs (Figure 1). For the multivariable MR adjusting for genetic confounding through BMI, estimated glomerular filtration rate, type 2 diabetes and lipid traits, direct effects of these exposures on risk of the respective CVD outcomes are presented in Supplementary Table 3. Considering SBP, the main IVW and sensitivity MR analyses all provided supporting evidence of a causal effect of serum urate (IVW estimate in SD units per 1-SD increase in genetically predicted urate 0.09, 95%CI 0.05-0.12, *P*=6×10-7; Figure 2). Scatter plots depicting the association of the instrument variants with serum urate and the respective outcomes are presented in Supplementary Figures 1-4.

**Figure 1.**
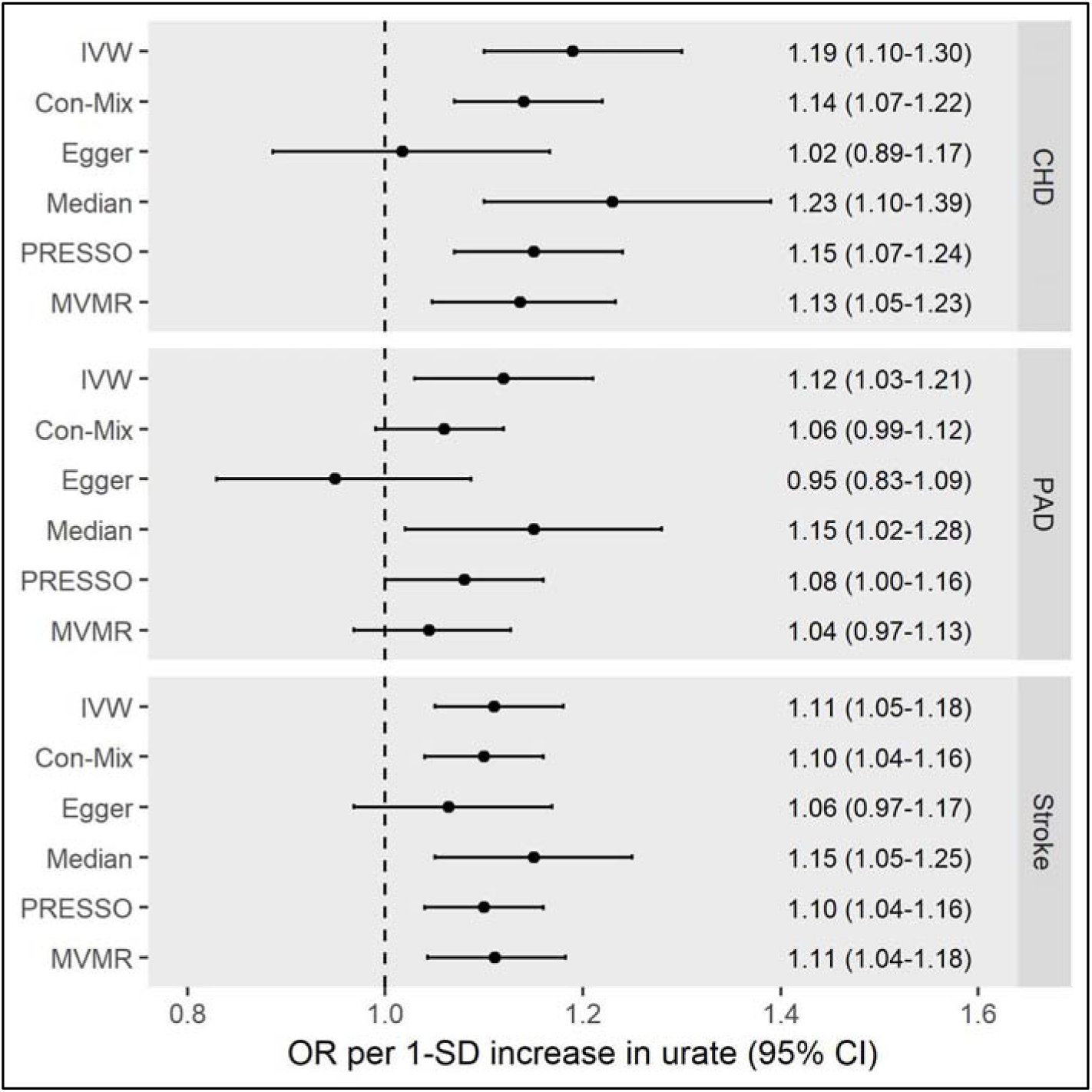
Mendelian randomization estimates for the effect of 1-standard deviation (SD) increase in genetically determined serum urate levels on risk of coronary heart disease (CHD), peripheral artery disease (PAD) and stroke. CI: confidence interval; Con-Mix: contamination-mixture; IVW: inverse-variance weighted; MVMR: multivariable Mendelian randomization (adjusting for genetic association of the instrument variants with body mass index, estimated glomerular filtration rate, type 2 diabetes mellitus, low-density lipoprotein cholesterol, high-density lipoprotein cholesterol and triglycerides); OR: odds ratio; PRESSO: pleiotropy residual sum and outlier. The outlier-corrected PRESSO results are presented (five outlier variants were identified for CHD, eight for PAD, and one for stroke). The Egger intercept was 0.005 (95% CI 0.001-0.009, *P*=0.004) for CHD, 0.005 (95% CI 0.001-0.009, *P*=0.003) for PAD and 0.001 (95% CI -0.001-0.003, *P*=0.24) for stroke.

**Figure 2.**
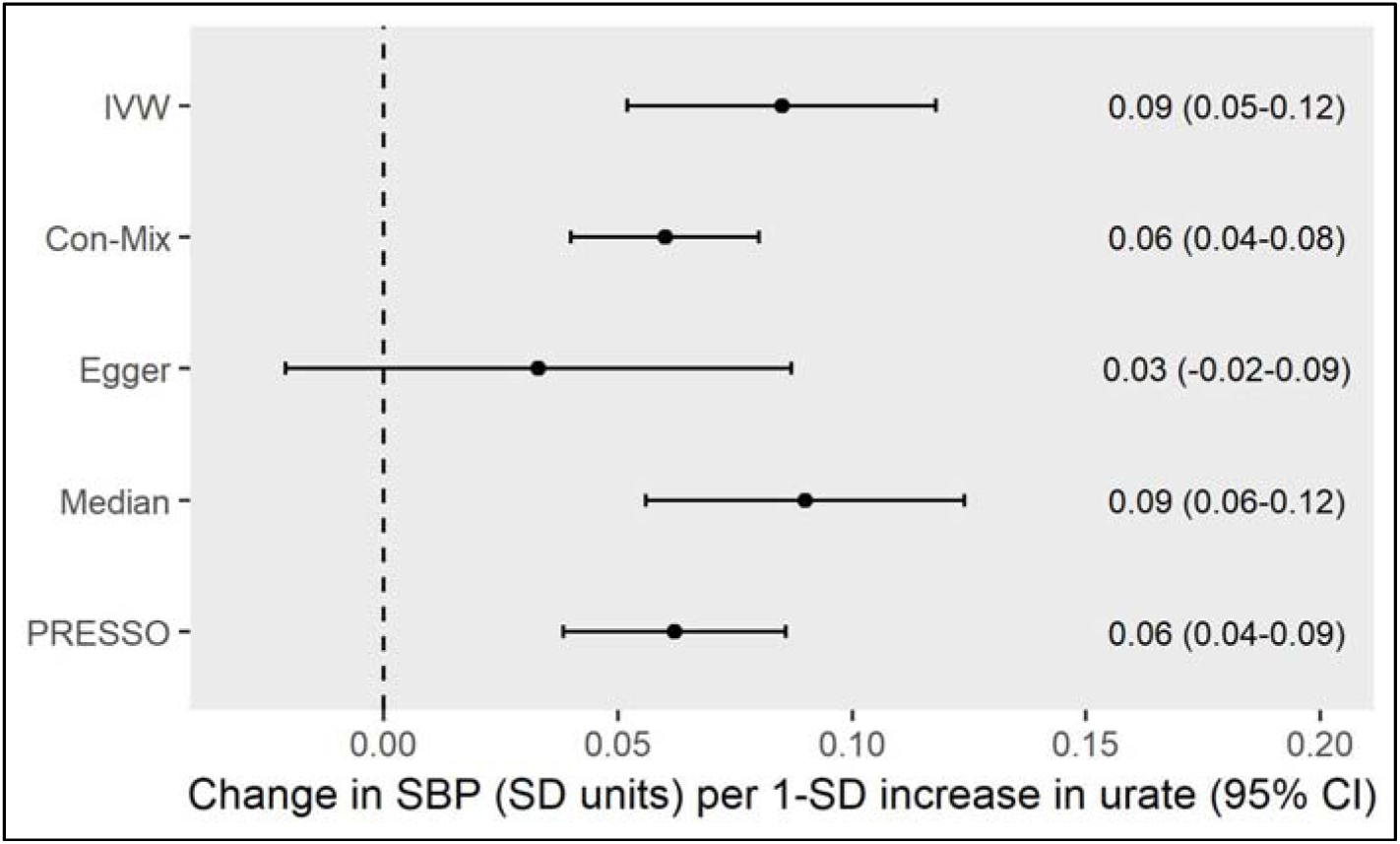
Mendelian randomization estimates for the effect of 1-standard deviation (SD) increase in genetically determined serum urate levels on systolic blood pressure. CI: confidence interval; Con-Mix: contamination-mixture; IVW: inverse-variance weighted; OR: odds ratio; PRESSO: pleiotropy residual sum and outlier. The outlier-corrected PRESSO results are presented (21 outlier variants were identified). The Egger intercept was 0.002 (95% CI 0.0004-0.003, *P*=0.02)

Performing multivariable MR to adjust for genetically predicted SBP showed attenuation of the urate effect estimates for the CVD outcomes as compared to the main IVW univariable MR (Figure 3), supporting that part of the effect of urate on these outcomes is mediated through SBP. Network MR mediation analysis quantified this as 29% (95%CI 9%-48%) for CHD, 44% (95%CI 5%-83%) for PAD and 45% (95%CI 14%-76%) for stroke. For CHD there remained evidence of a direct effect of urate even after adjusting for SBP (OR 1.13, 95%CI 1.03-1.23, *P*=0.01). In contrast for PAD and stroke, although the estimate for the direct effect of urate that is not mediated through SBP was positive, the confidence interval crossed the null and the results were not statistically significant (PAD OR 1.08, 95%CI 1.00-1.17, *P*=0.07; stroke OR 1.06, 95%CI 0.99-1.12, *P*=0.10). Direct effects of SBP on the outcomes after adjusting for genetically predicted serum urate are presented in Supplementary Table 4.

**Figure 3.**
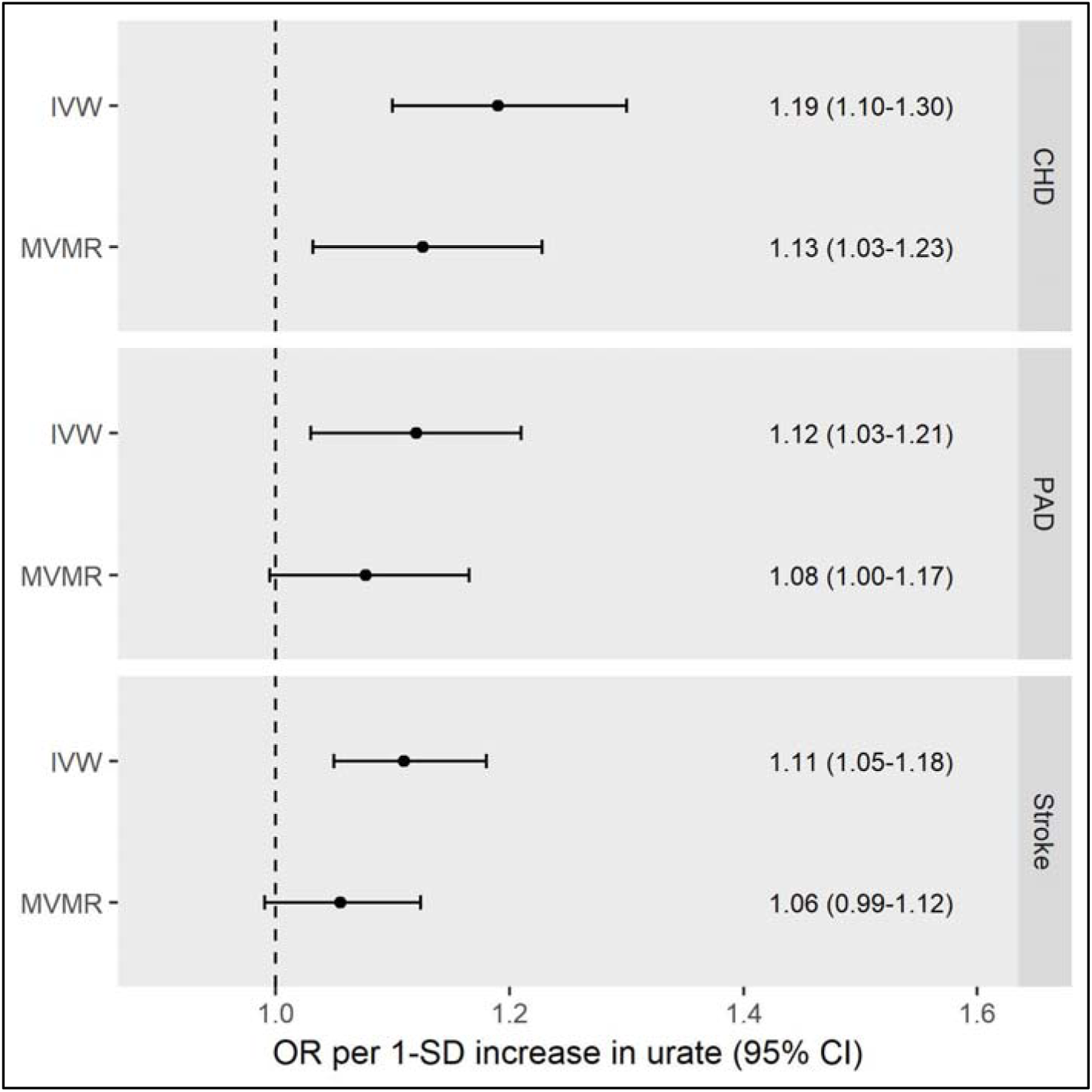
Inverse-variance weighted (IVW) and multivariable Mendelian randomization (MVMR) estimates for the effect of 1-standard deviation (SD) increase in genetically determined serum urate levels on risk of coronary heart disease (CHD), peripheral artery disease (PAD) and stroke. The MVMR analysis adjusts for the association of the genetic instruments with systolic blood pressure. CI: confidence interval; OR: odds ratio.

The results of the one-sample MR analysis were consistent with the above-mentioned two-sample MR analysis and are detailed in the Supplementary Results, Supplementary Tables 5-8 and Supplementary Figure 5.

**Figure 5.**
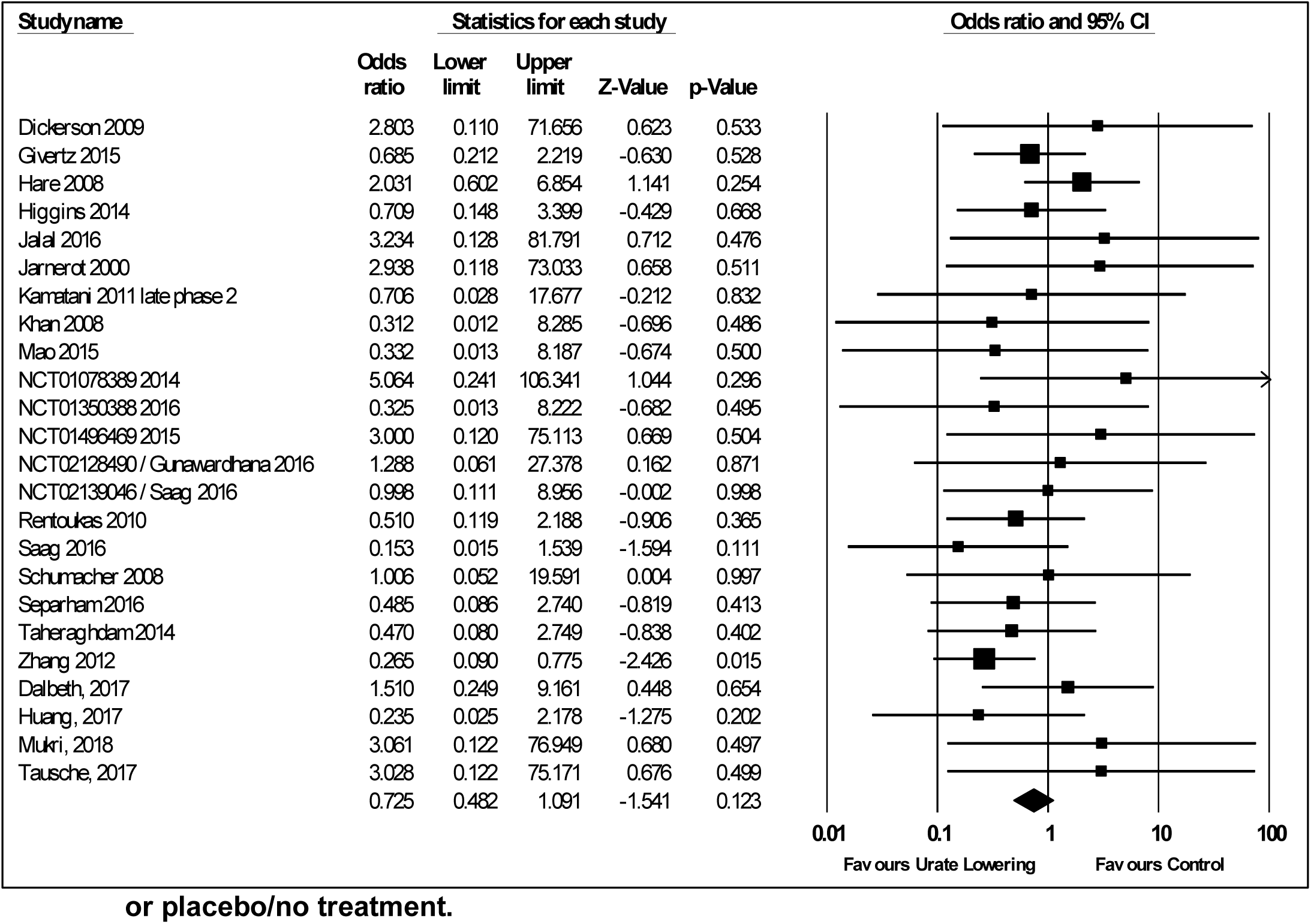
Forest plot of randomized controlled trial estimates for risk of major adverse cardiovascular events in all patients receiving urate-lowering therapy or placebo/no treatment. I^2^ heterogeneity statistic: 0%. CI: confidence interval.

### Systematic review and meta-analysis of randomized controlled trials

In total, 95 studies were eligible for the systematic review and meta-analysis. Regarding the updated searches (1 January 2016 to 30 September 2019), 5,353 records were screened, 30 full texts were assessed for eligibility and 13 studies were identified that fulfilled the eligibility criteria (Supplementary Figure 6). Characteristics of included studies that were identified in the updated search are presented in Supplementary Table 9.

**Figure 6.**
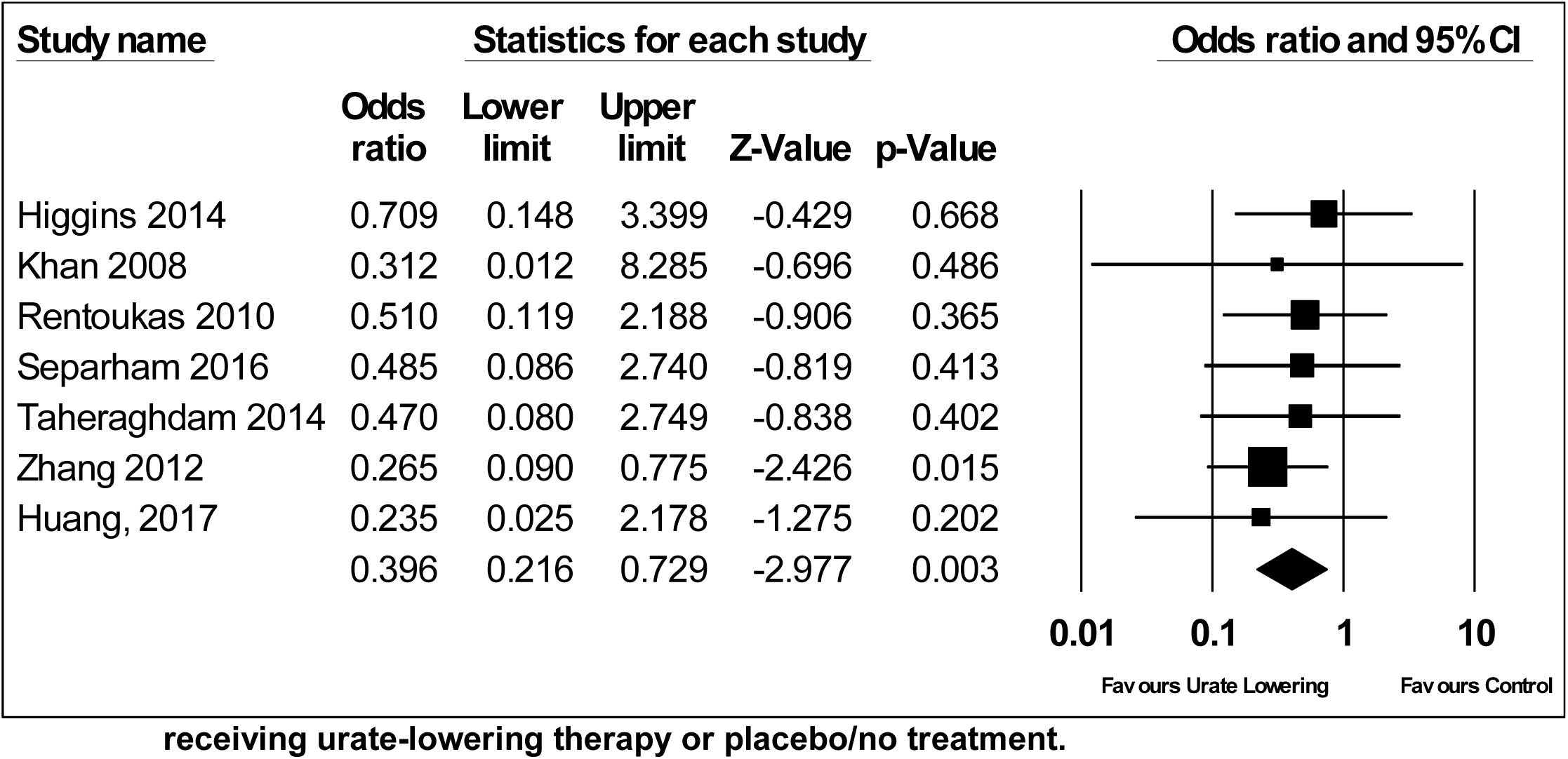
Forest plot of randomized controlled trial estimates for risk of major adverse cardiovascular events in patients with existing cardiovascular disease receiving urate-lowering therapy or placebo/no treatment. I^2^ heterogeneity statistic: 0%. CI: confidence interval.

The primary analysis of effects on SBP included 15 studies (Supplementary Figure 7). There was heterogeneity between these studies (I^2^ 89%), which was mostly attributable to one study considering renal dialysis patients (Supplementary Figure 7) (33). Excluding this study, subjects treated with urate-lowering therapy had greater reduction in SBP than subjects in the control group (mean difference in SBP -2.55, 95%CI -4.06 to -1.05, *P*=1×10^−3^, I^2^ 43%) (Figure 4). The analysis of MACE risk in all patients considered 88 studies, of which 24 had events (Figure 5), and the analysis of MACE risk in patients with prior cardiovascular events considered 10 studies, of which 7 had events (Figure 6). Urate-lowering therapy was not significantly associated with risk of MACE in all patients (OR 0.73, 95%CI 0.48 to 1.09, *P*=0.12, I^2^ 0%), but was significantly associated with reduction in risk of MACE in patients with a prior cardiovascular event (OR 0.40, 95%CI 0.22 to 0.73, *P*=3×10-3, I2 0%).

**Figure 4.**
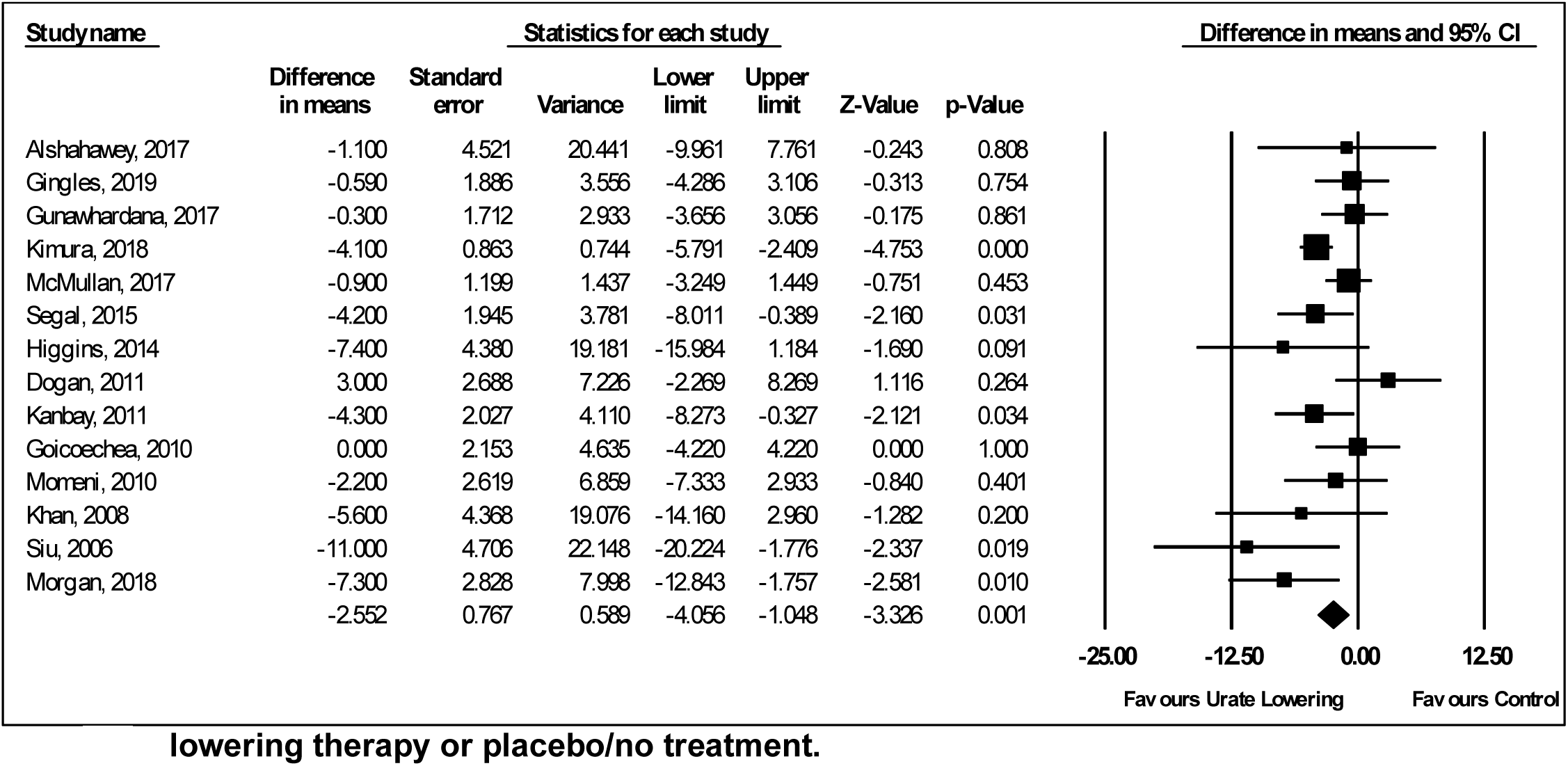
Forest plot of randomized controlled trial estimates for change in mean systolic blood pressure in non-renal dialysis patients receiving urate-lowering therapy or placebo/no treatment. I^2^ heterogeneity statistic: 43%. CI: confidence interval

Risk of bias was low in 23 of the 95 included studies (Supplementary Table 10). Risk of bias was low in 4 out of the 15 studies investigating effects on SBP, 20 out of the 24 studies investigating MACE in all patients, and 4 out of the 7 studies investigating MACE in patients with a prior cardiovascular event. In sensitivity analyses including only studies at low risk bias, urate lowering therapy was associated with greater SBP lowering compared to control (mean difference in SBP -2.89, 95%CI -5.37 to -0.41, *P*=0.02, I^2^ 49%; Supplementary Figure 8), but was not significantly associated with risk of MACE in all patients (OR 0.64, 95%CI 0.28 to 1.46, *P*=0.29, I^2^ 0%; Supplementary Figure 9) or in patients with a prior cardiovascular event (OR 0.59, 95% CI 0.18 to 1.91, *P*=0.38, I^2^ 0%; Supplementary Figure 10). Meta-regression analysis demonstrated no effect of baseline SBP on treatment response to urate lowering therapy (coefficient -0.01, 95% CI -0.19 to 0.17, *P*=0.92; Supplementary Figure 11).

## Discussion

We used a comprehensive framework of MR methodologies to perform detailed investigation into the association of genetically predicted serum urate with CVD outcomes, and replicated findings in the independent UK Biobank population. The MR analyses identified consistent evidence of an effect of higher genetically predicted serum urate levels on risk of CVD and SBP. Performing multivariable MR to adjust for genetic association with SBP attenuated the estimates for the CVD outcomes to support that at least some of the effect of urate may be mediated through raised blood pressure. Updated systematic review and meta-analysis of RCT data similarly showed a favorable effect of urate-lowering treatment on SBP, with some evidence to also support a protective effect on MACE risk in individuals with prior CVD, but not in all individuals.

Hyperuricemia has been postulated to cause endothelial dysfunction by increasing oxidative stress (1), and this could directly increase risk of CVD through effects on the vascular endothelium (1). In an animal model where hyperuricemia was induced using a uricase inhibitor, hypertension followed after three weeks while controls remained normotensive (5), thus representing a second mechanism by which urate can increase risk of CVD. The results of the Febuxostat and Allopurinol Streamlined Trial (FAST) and the Allopurinol and cardiovascular outcomes in patients with ischemic heart disease (ALL-HEART) trials will help provide further insight on potential re-purposing of existing urate-lowering agents for CVD prevention (34, 35). Allopurinol could represent an inexpensive, safe and well-tolerated drug for reducing cardiovascular risk (36).

Our current study has several strengths. We triangulate evidence across MR analyses and RCT data to provide consistent evidence for a role of serum urate in increasing SBP and potentially also increasing CVD risk. Our analyses identify a possible mediating role of SBP in the pathway from serum urate to CVD risk. In contrast, previous MR studies have considered associations of the genetic instruments for urate with blood pressure as representing genetic confounding (3, 9-11). Exclusion of such variants or adjustment for their genetic association with blood pressure traits, as was done in these studies (3, 9-11), would obscure any true causal effect of genetically predicted urate on cardiovascular disease risk. Our present MR analysis also incorporated more powerful instruments for serum urate than were available previously. The variants used as instruments in our one-sample MR analysis of UK Biobank participants explained approximately 7.7% of the variance in serum urate (4), in contrast to the 5.3% explained when selecting instrument variants from the previous largest published GWAS of serum urate (13). Similarly, our systematic review and meta-analysis of RCTs updated evidence from previous efforts to consider an additional 13 studies (7, 8).

Our work also has limitations. The MR approach makes a series of modelling assumptions, and in particular requires that the genetic variants used as instruments do not affect the considered outcomes through pathways that are independent of urate. While this can never be completely excluded, we performed a range of MR sensitivity analyses that make distinct assumptions on the presence of pleiotropic variants and generally found consistent estimates. The MR-Egger method generated wide 95%CIs in all analyses and was likely of limited reliability due to the strength of the urate instruments being correlated to their direct effects on the CVD outcomes under consideration (4, 37). The majority (71 out of 94) RCTs considered in our meta-analysis had a high or undetermined risk of bias and there was heterogeneity in the results of RCTs measuring change in SBP following urate-lowering therapy, possibly related to study design, in turn limiting the strength of conclusions that can be drawn. Finally, we investigated SBP in mediating the effect of urate on CVD outcomes, rather than diastolic blood pressure. These two traits are highly genetically and phenotypically correlated (17), and it follows that a similar mediating role may be found for diastolic blood pressure.

To summarize, we have found consistent MR and RCT evidence for an effect of higher serum urate levels on increasing SBP, with further evidence also supporting a potential effect on risk of CVD. High-quality trial data are now necessary to provide definitive evidence on the specific clinical contexts where urate-lowering may be of cardiovascular benefit, with some large-scale studies already underway (34, 35).

## Contributors

DG, JD, ACC, ET and IT designed the study. DG, XL, ACC, DD, AHA-R and MT-R had full access to the data and performed the analysis. All authors interpreted the results. DG, JD, ACC and IT drafted the manuscript. All authors critically revised the manuscript for intellectual content. All authors approved the submitted version and are accountable for the integrity of the work.

## Acknowledgements

The authors acknowledge the contributors of the data used in this work: CARDIoGRAMplusC4D, CKDGen, DIAGRAM, Genetic Investigation of ANthropometric Traits, Global Lipids Genetics Consortium, International Consortium for Blood Pressure, MEGASTROKE, Million Veterans Program and UK Biobank.

